# Optimizing long-term prevention of cardiovascular disease with reinforcement learning

**DOI:** 10.1101/2024.12.09.24318697

**Authors:** Yekai Zhou, Ruibang Luo, Joseph Edgar Blais, Kathryn Tan, David Lui, Kai Hang Yiu, Francisco Tsz Tsun Lai, Eric Yuk Fai Wan, CL Cheung, Ian CK Wong, Celine SL Chui

## Abstract

The prevention of chronic disease is a long-term combat with continual fine-tuning to adapt to the course of disease. Without comprehensive insights, prescriptions may prioritize short-term gains but deviate from trajectories toward long-term survival. Here we introduce Duramax, a fully evidence-based framework to optimize the dynamic preventive strategy in the long-term. This framework synchronizes reinforcement learning with real-world data modeling, leveraging the diverse treatment trajectories in electronic health records (EHR). In our study, Duramax learned from millions of treatment decisions of lipid-modifying drugs, becoming specialized in cardiovascular disease (CVD) prevention. The extensive volume of implicit knowledge Duramax harnessed far exceeded that of individual clinicians, resulting in superior performance. Specifically, when clinicians’ treatment decisions aligned with those suggested by Duramax, a reduction in CVD risk was observed. Moreover, post hoc analysis confirmed that Duramax’s decisions were transparent and reasonable. Our research showcases how tailored computational analysis on well-curated EHR can achieve high nuance in personalized disease prevention.

## Introduction

Early prevention is one of the most effective and cost-efficient approaches to reducing the global disease burden^1^. Effective prevention typically involves two steps: identifying patients who require preventive measures and individualizing follow-up preventive strategies for these patients^2^. With advancements in artificial intelligence (AI) for healthcare, numerous prognostic tools have been developed, offering higher accuracy and adaptability^3^. These tools have been extensively used to assist in identifying patients at elevated risk for early prevention^4^. However, there has been limited progress to assist the subsequent step, i.e., individualizing long-term dynamic preventive strategies.

For example, as the leading cause of death globally^1^, cardiovascular disease (CVD) often relies on lipid-modifying drugs (LMDs)^5^ for primary prevention. While clinical guidelines^6–8^ recommend risk prediction models^4,9–13^ to identify patients needing lipid control, there is currently no tool to assist clinicians in tailoring preventive strategies to these patients. This lack of support leads to significant variability in prevention strategies^14^, where the accumulation of suboptimal treatment decisions can eventually deviate patient trajectories from long-term prevention of CVD^15^.

Such a complex decision-making task holds promise from recent advancements in AI. Reinforcement learning (RL)^16^, a branch of AI, is able to learn and employ a strategy to make a sequence of decisions that maximize future reward^17^, analogous to clinician’s goal to adapt prescriptions to optimize patient survival^18^. As an emerging technique in recent years, RL has already shown strength in several healthcare scenarios^19–21^, particularly in short-term patient outcome optimization such as in inpatients^22^ and intensive care units^23^ settings.

Here we present Duramax, a data-driven framework based on RL to optimize the long-term dynamic preventive strategy. We employed Duramax to provide personalized LMD treatments for the primary prevention of CVD. Leveraging a comprehensive and high-quality time-series dataset from Hong Kong over the past two decades, we conducted extensive analyses to investigate the real-world dynamics of lipid responses to different LMDs. Building upon a practical model that captures lipid dynamics, we developed Duramax in a way that emphasized its interpretability and ensured the safety of its recommendations in a separate post hoc analysis. By leveraging RL techniques and real-world data modeling, Duramax showcases its proficiency in tailoring its recommendations to suit specific local healthcare settings. Through this customization, Duramax offers optimal prescriptions that demonstrate lower expected CVD risk compared to clinician practice, as demonstrated by the validation results.

## Results

### Dataset preparation

Our study leveraged the data source provided by the Hong Kong Hospital Authority (HA), the largest public healthcare provider responsible for capturing over 70% of all hospitalizations in Hong Kong for more than two decades^24^. The HA also provides outpatient services in primary, secondary, and tertiary settings. All medical records were linked with an anonymized unique patient identifier. We collected patient disease diagnoses, prescription records, clinical lab tests, and healthcare utilization data from the period spanning 2004 to 2019. From a pool of around one and a half million patients under primary prevention of CVD since 2024, we selected around one-third of patient trajectories with high completeness of lipid test and LMD prescription records. The inclusion and exclusion criteria are in Methods and Extended Data Fig. 1.

Specifically, the development cohort comprised 62,870 patients from Hong Kong Island, encompassing a total of 3,637,962 treatment months. Within this cohort, we identified 214 different types of lipid-modifying drugs and combinations, providing a rich selection pool for the RL agent to potentially choose from (Fig. 1a). Furthermore, the validation cohort consisted of 454,361 patients from Kowloon and New Territories, covering a total of 29,758,939 treatment months (Fig. 1d). The patient demographics and clinical characteristics are in Extended Data Table 1. This curation, to the best of our knowledge, represents one of the largest and the most comprehensive data sources to investigate LMD effectiveness in the primary prevention of CVD.

**Fig. 1.**
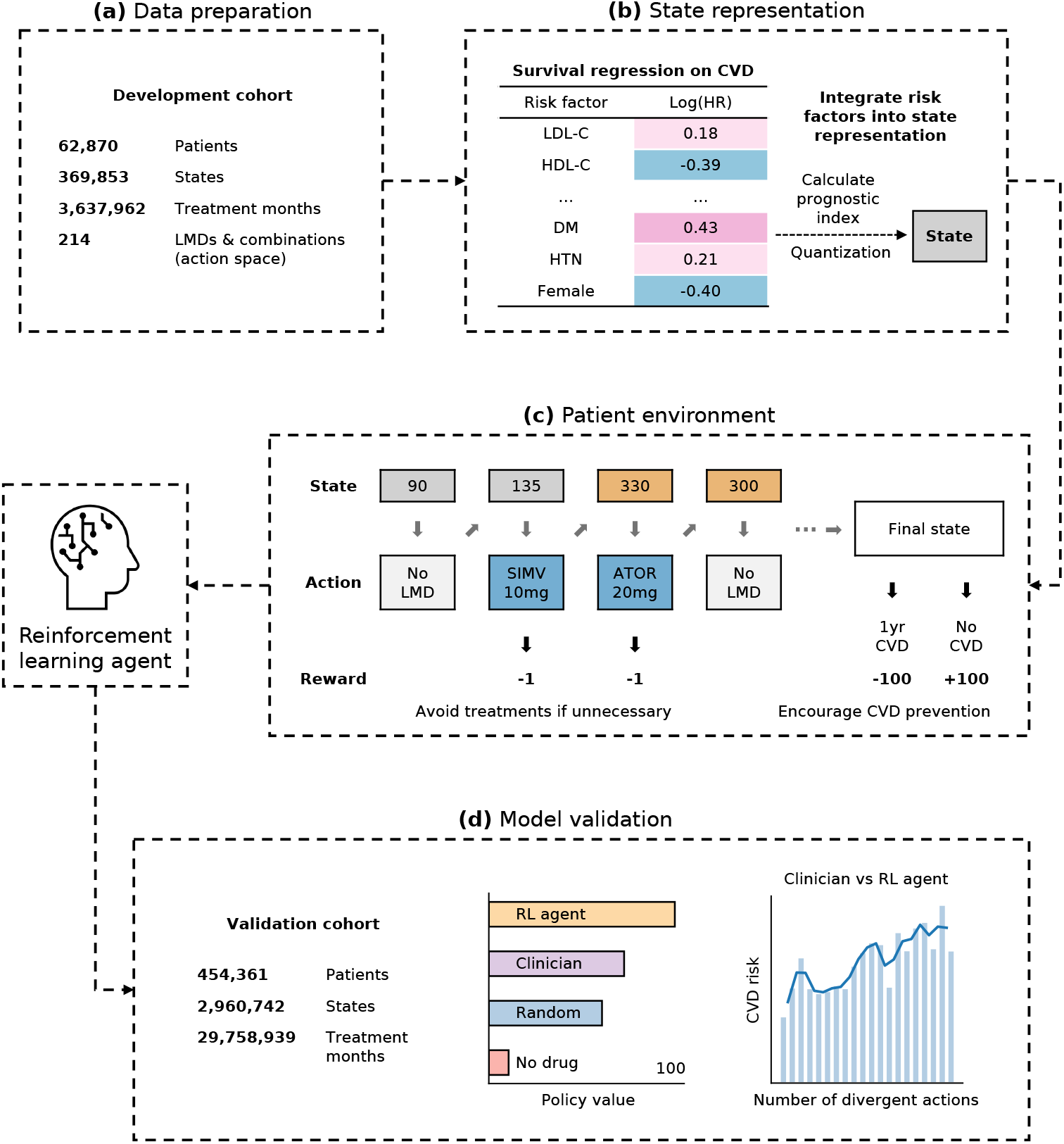
Overview of the study. Initially, a time series dataset of approximately 3.6 million treatment months was prepared for model development, encompassing an action space of 214 different types of lipid-modifying drugs (LMDs) and LMD combinations. Feature selection identified 10 key risk factors with quantified hazard ratios (HR) on cardiovascular disease (CVD) occurrence, which were integrated to represent patient states in a risk-based manner. The reward function penalizes unnecessary LMD treatments and final CVD occurrences, facilitating unconstrained exploration of optimal solution paths. Patient states, actions, and rewards were organized into transition matrix and fed into the reinforcement learning (RL) agent. Policy iteration, an offline model-based RL algorithm based on dynamic programming (DP), was chosen for its interpretability, stability, and guaranteed convergence to optimal solutions, making it particularly advantageous for high-risk applications like medicine. The RL agent was then validated using an independent cohort comprising approximately 30 million treatment months from 0.4 million patients. Validation results indicated that the RL agent exhibited superior performance compared to clinicians, with lower CVD risk observed as clinicians’ actions aligned more closely with the RL agent’s suggested actions.

### Modeling lipid dynamics

Developing a reliable RL agent for LMD prescriptions requires a comprehensive understanding of the complex lipid dynamics associated with different LMD use in real-world clinical settings. To fill this gap, we present a systematic analysis conducted on our development cohort. There are two primary objectives: 1) identify patterns and factors driving changes in lipid profiles, leading to a rational framework to represent complex patient trajectories observed in clinical practice, and 2) assess the long-term real-world efficacy of different LMD types, providing evidence that can guide treatment decisions for clinicians and the RL agent alike.

First, we analyzed the LDL-C dynamics over time (Fig. 2a). On average, patients on LMD treatment experienced an initial sharp reduction in LDL-C levels, with the median decreasing from 3.2 mmol/L to 2.2 mmol/L. Subsequently, a stable and gradual reduction rate was observed to help maintain consistently low LDL-C levels, resulting in a median LDL-C level of 2 mmol/L after a 60-month follow-up period. Patients not currently taking LMD (no LMD or stop LMD) showed a gradual reduction in LDL-C levels over time, with the median LDL-C level decreasing from approximately 3 mmol/L at the first visit to 2.8 mmol/L after a 60-month follow-up. This may be attributed to lifestyle modifications implemented during regular lipid testing.

**Fig. 2.**
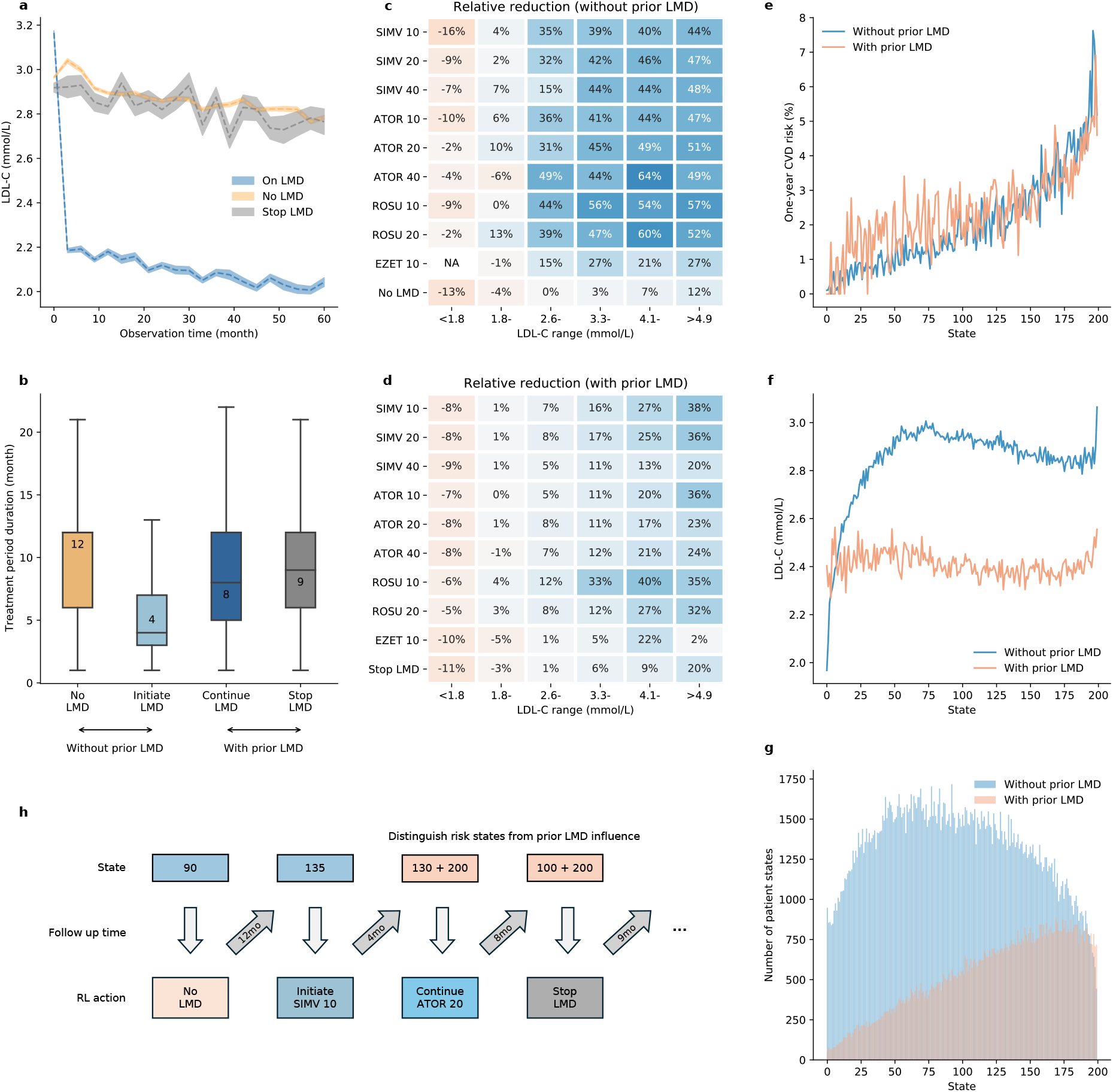
Analysis of lipid dynamics shaped RL environment design. Patients’ prior LMD usage had impact on future treatment effect on LDL-C reduction. As a result, the RL environment design distinguished four treatment categories: no LMD, stop LMD, continue LMD, and initiation of LMD. **(a)** The estimation of the LDL-C dynamics on three distinct scenarios: 1) patients who did not receive any LMD treatment (no LMD), 2) patients who initiated and continued LMD treatment (on LMD), and 3) patients who discontinued LMD treatment (stop LMD). Specifically, we selected patient trajectories that consistently followed the same actions during consecutive LDL-C tests over a period of up to 60 months. We calculated the population median of LDL-C levels for each quarter of observation time. The shaded area represents the standard error of the mean. **(b)** Distribution of treatment period duration across different treatment categories. The central line and the value indicate the median. The bottom and top edges of the box indicate the 25th and 75th percentiles, respectively. The whiskers extend to 1.5 times the interquartile range. **(c-d)** Median relative reduction of LDL-C based on different original LDL-C ranges, the most common 9 LMD types, and different treatment categories. Thicker shades of blue indicate a higher reduction rate, while thicker shades of orange indicate a higher increase rate. LDL-C levels were classified into six groups as indicated on the x-axis. "NA" indicates not applicable due to the absence of data points in that category. **(e-g)** Mean one-year CVD risk, mean LDL-C, and number of states across different risk states and different treatment categories. **(h)** Schematic illustration of the RL environment design. State numbers range from 0 to 199, representing risk states calculated for patients who had not taken LMDs before. After LMD initiation, state numbers are the original risk states plus 200, differentiating risk states influenced by prior LMD usage. Follow-up times were determined based on RL-suggested actions, using the median treatment period duration specific to each treatment category in (B). SIMV 10 = simvastatin 10mg. SIMV 20 = simvastatin 20mg. SIMV 40 = simvastatin 40mg. ATOR 10 = atorvastatin 10mg. ATOR 20 = atorvastatin 20mg. ATOR 40 = atorvastatin 40mg. ROSU 10 = rosuvastatin 10mg. ROSU 20 = rosuvastatin 20mg. EZET 10 = ezetimibe 10mg.

Consequently, we defined four distinct treatment categories between lipid tests: no LMD, initiate LMD, continue LMD, and stop LMD (details elaborated in Methods). The distribution of follow-up interval lengths also varied significantly among the four treatment categories (Fig. 2b). For patients who initiated LMD treatment, the next follow-up visit of lipid test had a median interval of 4 months. In contrast, patients not prescribed with LMD had a median follow-up interval of 12 months. This suggests that clinicians prioritize monitoring for patients starting LMD to assess the effectiveness whilst those who were not prescribed LMD were likely considered to have a lower CVD risk and require less frequent monitoring. These observations further distinguished the four treatment categories, where the variability in follow-up times indicates distinct clinical practices. It also highlights the importance of aligning our proposed RL agent with the clinician and healthcare system practice regarding follow-up intervals in real-world settings.

We further analyzed the effectiveness of commonly used first line and second line LMDs in reducing LDL-C levels in terms of different treatment categories. Besides, we examined the relative reduction separately for different ranges of baseline LDL-C levels, aligning with the clinical guideline thresholds. Detailed results can be found in Fig. 2c,d. It was confirmed that high-potency LMDs (e.g., rosuvastatin 10-20mg and atorvastatin 40mg) tend to yield a higher relative reduction compared to low-potency LMDs^25^. Interestingly, patients with higher baseline LDL-C levels generally experienced a higher reduction rate from LMD treatment, particularly when initiating therapy. Notably, even with low-potency LMDs (e.g., simvastatin 10-20mg), patients with elevated baseline LDL-C levels achieved significant reductions. For instance, patients with baseline LDL-C levels as high as 5 mmol/L who initiated treatment with a lowest potency LMD, simvastatin 10mg, can achieve an LDL-C level of approximately 2.8 mmol/L, which is considered acceptable for patients with modest baseline CVD risk. Conversely, patients with lower baseline LDL-C levels tended to have lower reduction rates from LMDs, particularly during drug continuation. For instance, in cases where patients had already received LMD treatment and achieved considerable LDL-C reduction, individuals with a higher baseline CVD risk may require further reductions. High-potency LMDs, particularly rosuvastatin 10mg, demonstrated effectiveness in these scenarios.

In summary, we observed that the use of LMD exhibited two distinct phases in LDL-C reduction: drug initiation and continuation. This observation motivated us to distinguish four treatment categories which capture the complexity of different patient trajectories. These treatment categories exhibited unique follow-up times and patterns of LDL-C reduction, indicating that they are independent drivers of patients’ lipid responses in addition to the efficacy of different LMDs. Apart from all the factors investigated, it was also important to treat patients’ baseline CVD risk as a separate factor from LDL-C levels when personalizing real-world prescriptions.

Consequently, for the rational design of a RL agent for LMD prescription, it becomes essential to address two key considerations: 1) informing the agent about the patient’s prior LMD usage, and 2) enabling the agent to comprehend the patient’s baseline CVD risk.

### Model design

Long-term CVD prevention can be abstracted as a Markov decision process (MDP) framework^17,18^. The goal is to develop a digital prescription guideline (policy) that recommends LMDs (actions) based on individual risk profiles (states). The objective is to continuously reassess these risk profiles and adapt the recommendations over time (sequential decision-making), aiming to minimize the long-term risk of CVD occurrence while avoiding too high a dose of LMDs to prevent dose-related adverse effects (rewards and penalties). We use RL to solve the formulated MDP. Starting from a random policy, The RL agent iteratives its policy by exploring if adjustments in the prescription strategies (action-state designations) can yield a better policy with higher expected rewards. The detailed settings of the MDP are illustrated in the Methods section.

Ideally, the MDP should succinctly capture the dynamics of patient responses while preserving the interpretability in each of its components. To this end, we presented patient state in a risk-based manner, where the contribution of each risk factor was explicitly quantized in the state number (Fig. 1b, details in Extended Data Fig. 2 and Methods). In such way, patient state served as a reliable indicator of CVD risk for both states with prior LMD usage and those without (Fig. 2e). Conversely, although patients on LMD generally had higher risk (Fig. 2g), these patients did not exhibit positive association between LDL-C and state compared to patients not on LMD (Fig. 2f). This observation further supported that drug continuation generally had lower LDL-C reduction rate (Fig. 2a), which motivated us to encapsulate the original risk-based patient state with the additional information of prior LMD usage (details in Methods). Such a design also distinguishes four treatment types (no LMD, initiate LMD, continue LMD, and stop LMD) in the action space that we previously confirmed essential. The resulting patient trajectory with sequences of successive states and actions is illustrated in Fig. 2h.

The primary objective of CVD prevention is to promote effective prevention while minimizing unnecessary treatments. We translated this objective into the design of our reward function. Specifically, patients either receive a high penalty if they experience a CVD event within a year or receive a high reward if they remain event-free in the final state. Additionally, small penalties are imposed for each LMD taken (Fig. 1c). This design allows for comprehensive control over the ultimate outcome and internal side effects, without imposing excessive constraints, and eventually empowers the RL agent to explore the optimal solution paths to a full extent. More details about the reward function design can be found at Methods.

We employed policy iteration^23,26^, an offline, model-based, and dynamic programming (DP) RL method. For healthcare settings, offline RL is required as it can be trained exclusively from historical data^27^, eliminating the need for real-time exploration of random prescriptions on patients required by online RL algorithms. Furthermore, in contrast to model-free techniques, model-based methods can offer the rationale behind prescriptions^28^, a feature highly valued by clinicians for ensuring safety^22^. Additionally, the guaranteed convergence to optimal solutions inherent in DP methods renders them particularly advantageous for high-risk applications like medicine. We trained the RL agent and named it Duramax and described with details about the setting and training of the RL algorithm in Methods.

### Interpretation of the RL policy

We summarized the logic that Duramax (the RL agent) recommended patient treatments using data from the development cohort (Fig. 3). Duramax primarily recommended three types of actions for initiating LMD therapy: simvastatin 10 mg, simvastatin 20 mg, or no initiation of LMDs if the patient’s risk is deemed modest by Duramax. For follow-up treatment decisions, Duramax suggested a wider range of options and higher intensity of LMDs based on patient state, including simvastatin 10-40 mg, atorvastatin 10-20 mg, and rosuvastatin 10 mg. Duramax also suggested to discontinue treatment if it feels safe about the patient’s current risk profile.

**Fig. 3.**
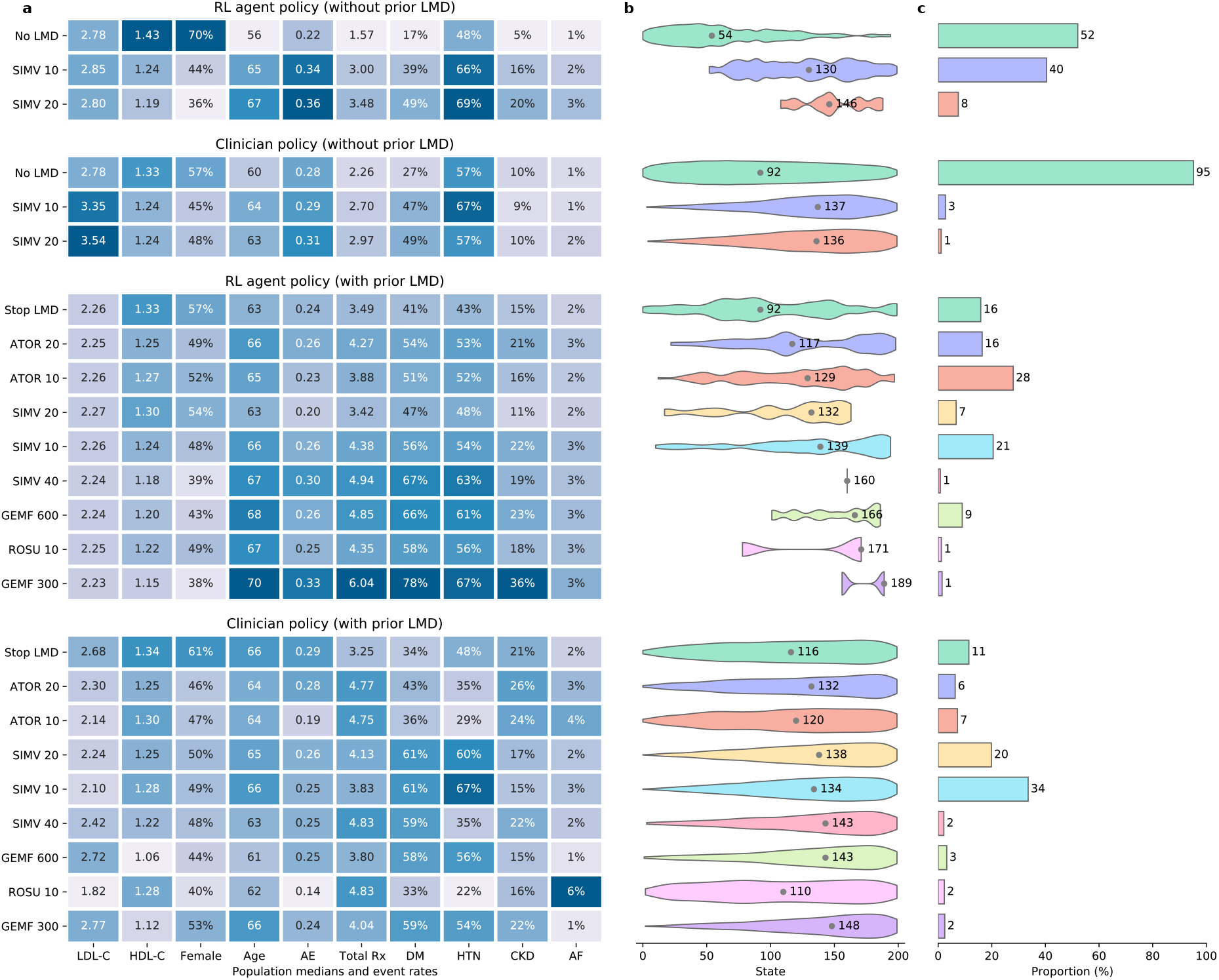
The RL agent policy demonstrated a comprehensive perspective on risk profile in specifying LMDs for patients compared to the clinician policy. **(a)** The population median values for continuous risk factors and event rates for dichotomized risk factors, **(b)** the distribution of state number in violin plots, and **(c)** the proportion of the target patients for whom the RL agent and clinician made different prescription choices. The treatments on the y-axis are sorted based on the median of the risk clusters in the RL agent policy. The medians in the violin plots are indicated by grey dots. LDL-C = low-density lipoprotein cholesterol. HDL-C = high-density lipoprotein cholesterol. DM = diabetes mellitus. AE = Accident and emergency visits within the last one year. Total Rx = concurrent medication records the number of drugs prescribed within one month. CKD = chronic kidney disease. AF = atrial fibrillation. HTN = hypertension. GEMF 300 = gemfibrozil 300mg. GEMF 600 = gemfibrozil 600mg. GEMF 300 and GEMF 600 were prescribed twice a day.

Interestingly, gemfibrozil 300-600mg was frequently suggested by Duramax.

One major advantage of model-based RL is the transparent decision-making process. We visualized the decision-making process and plotted the average decision threshold that influenced the agent’s choices (Fig. 3a). Comparisons were made between the Duramax’s decisions and those made by clinicians, considering population median and event rates. In summary, Duramax demonstrated higher specificity and a more comprehensive perspective on patient risk profiles compared to clinician decisions in prescribing LMDs. The clinician’s treatment decision primarily relied on LDL-C levels, whereas Duramax considered ten risk factors, resulting in a grading relationship among them. This comprehensive evaluation allowed Duramax to exhibit better specificity for patients with different states, and CVD risk alike (Fig. 3b). Consequently, the target patient group differed in clinician policy and the RL policy (Fig. 3c).

The RL agent suggested initiating treatment with lower-potency statins, such as simvastatin 10- 20 mg, aligning with our previous conclusion that even low-potency statins can have good treatment effect when initiated (Fig. 2c). For patients in the model development cohort being considered of whether initiating LMD therapy or not, only 4% of patients received prescription from clinicians, while Duramax suggested that 48% of them should initiate. However, the low proportion of actual statin takers might stem from patient concerns regarding side effects.

Regarding LMD continuation, atorvastatin 10-20 mg was more frequently prescribed by Duramax (44%) compared to the clinician policy (13%), whereas simvastatin 10-20 mg was less commonly prescribed by Duramax (28% vs. 54%). Notably, fibrates were used more frequently for very high-risk patients as suggested by Duramax (10% vs. 5%), with a higher median risk state of 189 compared to 148 in the clinician policy. The RL agent’s rationale behind these recommendations may be attributed to the multiple comorbidities present in high-risk patients and the limited room for further LDL reduction given their already low LDL-C levels. In this context, incorporating HDL-C may be a sensible approach.

### Model validation

We conducted model validation using an independent large-scale time series dataset from the Kowloon and New Territories cohort, comprising half a million patients and spanning 30 million treatment months with 3 million intermediate states. The validation results, illustrated in Fig. 1d and further elaborated in Fig. 4, exhibited excellent performance of the model. Specifically, Duramax exhibits superior accuracy in prescribing specific LMDs and determining the optimal timing for initiating, switching, or stopping LMD therapy.

**Fig. 4.**
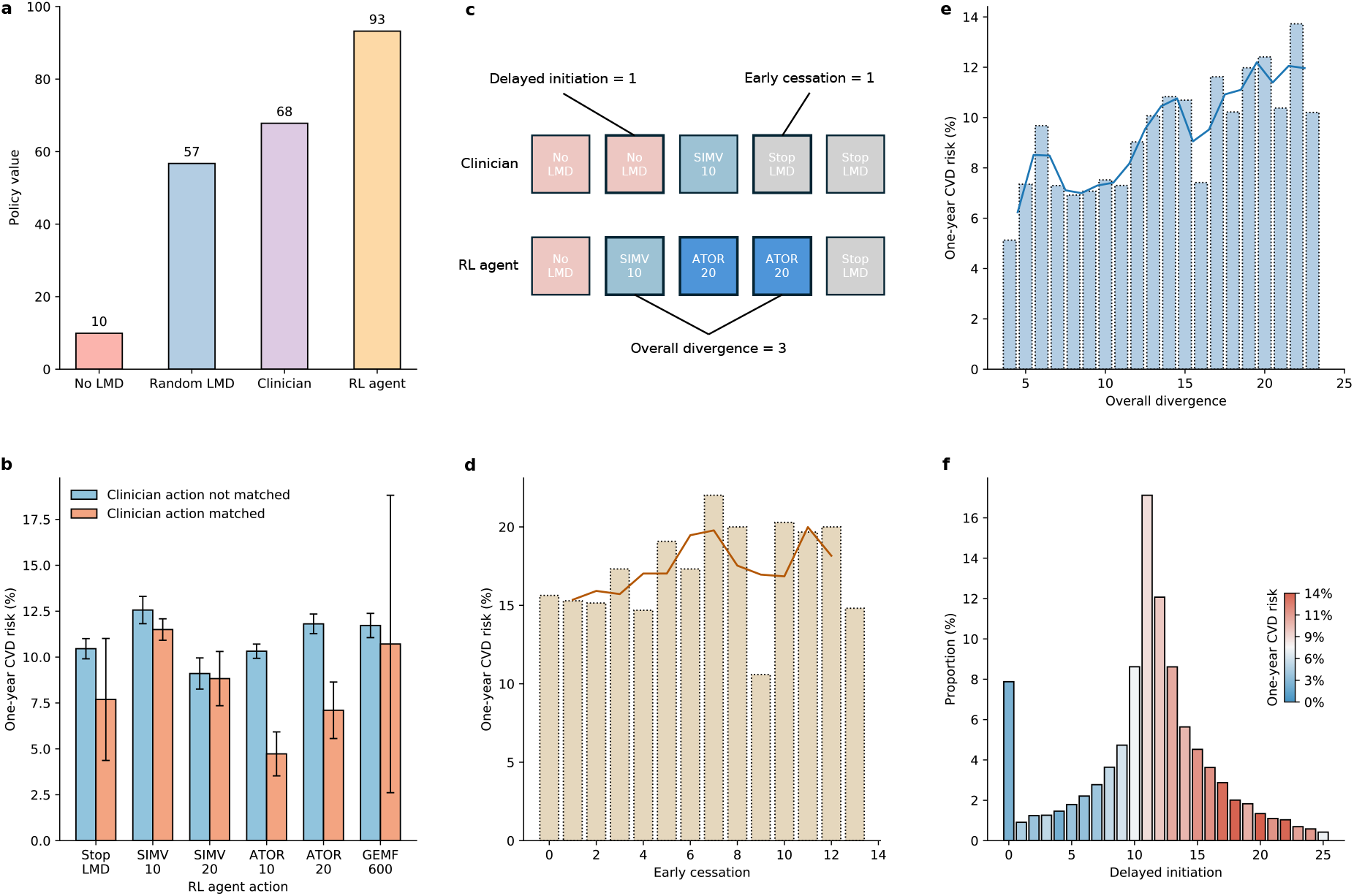
Model validation. The RL agent demonstrated superior accuracy in prescribing specific lipid-modifying drugs (LMDs) and determining the optimal time to initiate LMDs or stop LMDs. **(a)** The RL agent’s policy value exceeded the clinician’s policy as estimated by importance sampling. The clinician policy value was represented by the 95% upper bound, while the RL policy, random LMD policy, and no LMD policy were represented by the 95% lower bound, using bootstrapping with 1,000 resampling. **(b)** Patients whose last state matched the action suggested by the RL agent exhibited lower CVD risk. Actions with a minimum of 50 matched cases were selected for comparison to ensure statistical reliability. The error bars represent the 95% confidence interval. **(c-f)** The more actions the clinician aligned with the RL suggestion, the lower the CVD risk is. Specifically, higher CVD risk was observed in patients with higher early cessation, delayed initiation, and overall divergence. Only 8% of patients did not delay LMD treatment according to the RL agent’s estimation. The darkened lines in **(d-e)** represent smoothed results derived from the original consecutive bars. The unit in the x axis is the number of actions.

To evaluate the model’s performance, we first performed a gold standard check called off-policy evaluation using importance sampling (Fig. 4a, details in Methods). The estimated value of the RL policy (93) surpassed that of the clinician policy (68), where higher policy value is observed to be associated with reduced CVD risk (Extended Data Fig. 3). Additionally, two comparison policies, random drug policy (57) and no drug policy (10), had lower values than the clinician policy. Furthermore, we examined the prescription differences between the RL agent and clinicians. We observed that patients who received treatments suggested by the RL agent had the lowest CVD rate. We delved into the correctness of each specific action proposed by the RL

agent (Fig. 4b). For each type of action, we discovered that when the clinician’s action aligned with the RL agent’s recommendation, it resulted in lower CVD risk compared to misalignment. Notably, the greater the alignment between the clinician and RL agent actions, the lower the CVD risk for the patient (Fig. 4e). The RL agent also accurately identified the optimal time for patients to discontinue LMD therapy (Fig. 4d) and initiate LMD therapy (Fig. 4f). This capability makes the RL agent highly suitable for preliminary screening of patient groups eligible for primary prevention. It was estimated that only 8% of patients not delaying LMD treatment according to the RL agent’s estimation.

## Discussion

Approximately 50 years ago, the Framingham Heart Study investigators developed multivariable CVD risk prediction equations to assist in personalizing CVD treatment planning^13^. Since then, there have been continuous efforts to advance risk prediction models^4,9–12,29–31^ and refine clinical guidelines^6,7^ for improved nuance. Our study made a significant contribution to the field by creating the first fully data-driven evidence-based guideline with high nuance to individualize long-term treatment for CVD prevention. Through comprehensive validation, our RL agent demonstrated superior efficacy in reducing CVD risk compared to clinician consensus. In leveraging Duramax, we address the uneven distribution of healthcare resources by tapping into the wealth of treatment experiences documented in EHR. Our aim is to bolster healthcare support, particularly in regions where resources are scarce. Duramax can be use by junior clinicians during follow-up visits. By harnessing the insights provided by Duramax, these clinicians can better understand patients’ health trajectories and make informed decisions based on evidence-based suggestions generated by the system. Moreover, pharmacists and nurses can seamlessly integrate Duramax into their monitoring protocols. This integration allows them to track patients’ progress more effectively and intervene promptly when necessary. We envision that even a modest reduction in CVD risk by partially considering the suggestions from the RL agent has the potential to save millions of lives annually around the world.

While RL has been applied in various medical contexts^32^, our study significantly advances its utilization in long-term chronic disease prevention. This research contributes to the expanding body of knowledge on AI applications in preventive medicine and sheds light on the computational management of other chronic diseases, such as diabetes, hypertension, and obesity. Looking ahead, a promising future application lies in the development of personalized digital twins that optimize long-term health outcomes across multiple chronic disease domains. Extended from Duramax, the complex digital twin can monitor simultaneous the variations of key index tests e.g. glucose, blood pressure, BMI, as well as the lipid levels. The complex RL agent can monitor the health status and suggest preventive suggestions by incorporating a more multifaceted reward function design that penalizes the occurrence of multiple diseases and considering treatment options such as metformin and ACE inhibitors. This direction would contribute to the advancement of holistic management for chronic diseases but also warrant a more sophisticated curated EHR data that aligns enough clinical lab tests for monitoring.

To simulate a realistic lipid dynamic model for hosting the RL agent, our study undertook one of the most extensive and comprehensive investigations to date, exploring the intricate relationship between lipid levels, drug interventions, and screening practices in real clinical settings. Unlike previous studies in this field that primarily focused on randomized controlled trials^33–37^ of single drugs in small populations with similar risk profile, our study examines lipid dynamics across various LMDs using a large-scale, long-term observational approach. By adopting this approach, we were able to uncover significant patterns in lipid metabolism and treatment response, which bridged the gap between controlled trials and real-world lipid management.

This comprehensive analysis provided a practical model for understanding lipid dynamics under the influence of different LMD types in real-world scenarios. Importantly, it offers clinicians and researchers a valuable resource to comprehend the dynamics and expected treatment efficacy over time, enabling informed decision-making when formulating lipid management strategies.

Our research presents a practical approach to develop AI-based complex decision support systems using EHRs, which achieves both interpretability and accuracy in its design. In recent years, deep learning methods have gained prominence in healthcare AI applications^38–40^, being positioned as end-to-end solutions where the model takes in the original input and produces the output through a series of complex computations^41^. While this approach has shown comparable performance, it often lacks interpretability^41^. In medical applications, understanding the decision-making process is as important as the outcome itself, particularly in scenarios without prior knowledge^42^. In contrast, we adopted a combination of classical approach to ensure the controllability of the intermediate steps. We employed a risk-based state representation and a model-based dynamic programming framework, enabling the development of transparent and interpretable AI solutions for sequential decision-making tasks. Furthermore, it turned out that the accurate representation of the patient’s dynamic lipid environment further facilitated the exceling performance of our agent. Looking forward, we identify several promising avenues for future research. These include the collection and integration of informative yet unstructured features, such as genomic data, medical imaging, or clinical free-text notes. By incorporating these elements into our state definition, we anticipate a significant enhancement in the model’s sensitivity and overall performance. In such scenarios, a separate deep learning component would be necessary to process and interpret these unstructured data sources^41^, while preserving the interpretability of the core decision-making process.

Our study presents a fully evidence-based approach to clinical decision-making that can be integrated into routine patient care through multiple avenues. One primary application is the incorporation of our RL agent into clinical guidelines as a reference tool for clinicians, providing evidence-based recommendations to support decision-making processes. Looking ahead, we envision the potential for enhanced human-AI interaction within this framework^43^. In this scenario, the RL agent would offer an initial treatment recommendation, which the clinician could then review and comment on. The RL model would subsequently refine its recommendation based on clinician’s comments, creating a dynamic and responsive system. This interactive approach holds significant promise, particularly if coupled with a large corpus of clinical free-text notes as a training set. By utilizing advanced techniques in deep learning, such as sophisticated language models, there is potential to enhance the system’s ability to interpret and respond to nuanced clinical feedback. Furthermore, our approach introduces a novel method for screening potential candidates for primary CVD prevention. Unlike traditional methods that rely on predefined thresholds^6,7^, our analysis is entirely based on long-term CVD prevention outcomes. This data-driven screening approach has the potential to identify at-risk individuals who might be overlooked by conventional criteria, thereby improving preventive care strategies. By providing these innovative tools and insights, our research aims to support clinicians in making more informed, personalized decisions while maintaining the critical role of human expertise in patient care. As these methods are refined and validated in clinical settings, they have the potential to significantly enhance the precision and effectiveness of CVD prevention and management strategies.

The study would benefit from international validation. Although the model shows strong performance within our study population in Hong Kong, its applicability to diverse international settings remains to be established. However, the collection of data with enough granularity, dimensionality, and quantity for validation from different countries and healthcare systems would be highly challenging. If applied internationally, the model would likely require recalibration to account for differences in prescription patterns, healthcare delivery systems, and population characteristics across different regions. Before widespread adoption can be considered, prospective clinical trials are essential.

## Methods

### Study Design and Participants

This study included patients who had utilized public healthcare services provided by the Hong Kong Hospital Authority (HA) since 2004. HA is the largest public healthcare provider in Hong Kong, offering government-subsidized primary, secondary, and tertiary care to all residents. It accounts for over 70% of all hospitalizations in Hong Kong^24^. Previous research has confirmed the reliability of the HA’s data source which has been extensively used in multinational collaborative studies^44^, including research on CVD and CVD drug studies^45^, with a positive predictive value of 85% for myocardial infarction and 91% for stroke^46^.

Two patient cohorts were identified based on their primary location of residence in Hong Kong: Hong Kong Island (Hong Kong West Cluster, HKWC) and Kowloon and New Territories. The Hong Kong Island (HKWC) cohort was utilized for model development, while the Kowloon and New Territories cohort served for model validation, ensuring there was no overlap between the development and validation groups. Specifically, the Hong Kong Island (Hong Kong West Cluster) cohort consisted of patients aged 18 or above who had undergone a lipid test at a hospital within the Hong Kong West Cluster between January 1, 2004, and December 31, 2019, as identified by the Hospital Authority. The Kowloon and New Territories cohort included patients aged 35 or above whose blood pressure was recorded in the Hospital Authority’s database between January 1, 2005, and December 31, 2019. Patients who predominantly sought healthcare on Hong Kong Island and those without a lipid test record during the study period were excluded. The cohort entry date was defined as the date of their first lipid test in any inpatient or outpatient setting since 2004. Patients were censored at the earliest occurrence of the first recorded CVD diagnosis, registered death, or the study’s end date (December 31, 2019). Patients who experienced a CVD event before the first lipid test or who died on the same day as the test were excluded from the cohort. The primary outcome was the initial diagnosis of CVD, as defined by the International Classification of Diseases, Ninth Revision, Clinical Modification (ICD-9-CM) codes. The outcome was a composite measure encompassing coronary heart disease, ischemic or hemorrhagic stroke, peripheral artery disease, and congestive heart failure (see Supplementary Table 4).

### Patient trajectory selection and formalization

To formalize patient trajectories, we defined states as the time steps of each lipid test and actions as the choice of LMD prescription between states. The trajectory consisted of repeated state-action pairs until reaching the cohort end date, with a final state indicating the occurrence of CVD within one year after the last state.

**State.** We selected representative real-world patient trajectories by applying a filtration process.

(1) We excluded patients with fewer than two lipid test records during the study period to ensure consecutive trajectories. (2) Considering clinical guidelines and common practice, we excluded trajectories with visit intervals of less than one month or more than two years to align with real-world reliability. (3) Trajectories with incomplete lipid profiles (missing LDL-C, HDL-C, or triglyceride measurements) were excluded. Each patient state included a risk profile comprising 90 features, such as disease history, laboratory test results, healthcare utilization, and medication count. Disease history encompassed any previous diseases recorded before the state, identified using ICD-9-CM codes (refer to Supplementary Table 5 for details). Laboratory test results were obtained on the same date as the state. Healthcare utilization was determined by the number of visits within one year prior to the state’s date. Medication count referred to the number of different drugs with different British National Formulary (BNF) codes prescribed within one month prior to the state’s date (refer to Supplementary Table 6 for different drugs identified).

**Action.** Representing the actions, which involve the specific LMDs or combinations of LMDs taken by patients during each interval between two consecutive states, poses significant challenges. The task becomes even more demanding when attempting to identify a series of actions from a sequence of LMD records associated with lipid tests, as the prescribed medications and laboratory records often do not align perfectly. A typical scenario involves lipid tests occurring at the 0th, 3rd, 6th, and 18th months of a patient’s trajectory, while a particular LMD is prescribed from the 3rd to the 9th month. In this case, the action for the first interval (0-3 months) is clear, indicating no LMD was taken. The action for the second interval (3-6 months) is also evident, representing the specific LMD prescribed during that period. However, the third interval (6-18 months) presents ambiguity, as the prescription only covers (9 - 6) / (18 - 6) = 25% of the interval. Determining whether to consider the third action as a continuation or discontinuation of the LMD becomes uncertain, and deciding whether to include trajectories with such ambiguous actions poses a challenging choice. The complexity further escalates when multiple types of LMDs need to be considered simultaneously.

To prioritize representative and high-confidence trajectories, we implemented an empirical strategy consisting of the following steps:

**Calculating LMD Coverage.** We calculated the coverage of LMDs for each interval between two consecutive lipid tests. For instance, if a patient had a total prescription of simvastatin 10mg covering half of a six-month interval, the coverage for simvastatin 10mg would be 50%. We performed this calculation for multiple types of LMDs recorded in the database, considering each interval within the patient trajectory.

**Excluding Ambiguous Trajectories.** To ensure the quality of included trajectories, we considered any trajectory that had intervals with LMD coverage ranging from 1% to 50% as ambiguous in terms of drug continuation and discontinuation. Consequently, we excluded the entire trajectory if any intervals fell within this coverage range. If an interval had multiple LMDs prescribed with coverage above 50%, it was considered a combination of LMDs. Thus, we only considered trajectories that were unambiguous in terms of no drug, drug initiation, continuation, and discontinuation throughout their entire trajectory.

**Handling False Combination of LMDs.** Consecutive intervals might exhibit false combinations of LMDs, e.g., representing early transitions between drugs where the prior prescription was long enough to cover the next interval by more than half. To mitigate these artifacts, we examined the prescriptions in the last interval of each patient trajectory, which are generally more stable as they approach the end. The set of prescriptions in the last interval was considered the final set of actions. We removed patient trajectories that had prescriptions not matching the defined set of actions.

By following this approach, we were able to define patient actions in terms of LMD types, ensuring representative and reliable trajectories.

### Risk-based state representation

In order to incorporate the overall CVD risk level into each state, we aimed to quantify the contribution of individual features within the states. To achieve this, we performed survival analysis on the development cohort, considering the start time of their last state until the observation of CVD occurrence. Initially, we conducted a robust feature selection process to identify significant features associated with CVD occurrence^30^. For statistical reliability and clinical relevance, we selected features without missing values (e.g., clinical laboratory tests) and an event rate above 1% (e.g., disease and medication history). The Cox proportional hazards model (CPH)^47^ with least absolute shrinkage and selection operator (LASSO) regularization^48^ was employed to identify statistically significant features (p value < 0.05). The CPH model is widely used for survival analysis, and its regression coefficients can be interpreted as hazard ratios, facilitating better decision-making by clinicians. LASSO is a robust feature selection method that chooses a representative and independent set of features, ensuring reliability for downstream manual prioritization. The final set of features was also determined based on current clinical evidence to ensure comprehensiveness and relevance to CVD prognosis. Subsequently, we applied CPH with ridge regularization on the final feature set to quantify the contribution of each identified feature to CVD occurrence. Ridge regularization, a widely used stabilizer of regression coefficients, provided reliable estimates of hazard ratios for the risk variables. The contribution of each feature was represented as the natural logarithm of the hazard ratio. The calculation details for the state number are provided in Extended Data Fig. 2. The feature selection results are presented in Supplementary Tables 1-3. To assess the overall risk, we calculated a prognostic index (PI) for each patient by summing the contributions of individual features. The PI allowed us to unify the overall risk of different patients on the same scale. Next, we sorted the PIs of patients in the development cohort and divided them proportionally into clusters, with each cluster corresponding to a state number. Consequently, the state number now incorporates information about CVD risk, and its increase reflects an increasing CVD risk in an interpretable and transparent manner. It is important to note that the specific number of clusters (i.e., states) was determined through manual prioritization based on qualitative evaluation of the RL policy decision boundary during model development. We added 200 to the state number to indicate states after the initiation of LMDs (ranging from 200 to 399), distinguishing them from states without prior LMD usage (ranging from 0 to 199). This state representation, which accurately captures the patient’s overall CVD risk, enables the RL agent to make more informed decisions. Furthermore, an added advantage of this state representation is that actions considered within the same state number pool share a similar baseline CVD risk, which helps mitigate selection bias. Selection bias, a significant concern in retrospective studies, occurs when higher-risk patients are more likely to be prescribed high-intensity LMDs and may still experience a higher risk of CVD compared to low-risk patients using low-intensity LMDs. This approach also facilitated a direct comparison of the safety line for LDL-C. For example, by accounting for the coefficients of 0.43 for diabetes and 0.18 for LDL-C per unit increase, we can determine that patients with diabetes and an LDL-C level of 3 mmol/L have an approximate CVD risk similar to patients without diabetes but with an LDL-C level of 5 mmol/L.

### Formalization of the computational model

We formulated the patient trajectory and treatment decision-making process as a Markov decision process (MDP)^17^. The MDP was defined by the tuple [S, A, T, R, γ], where:

- S is a finite set of states representing the risk states of patients during their healthcare visits for lipid tests (as described in the previous section).
- A is the finite set of available actions representing the chosen LMD and LMD combinations (as described in the previous section).
- T(s’ | s, a) is the transition matrix, which determines the probability of transitioning from state s at time t to state s’ at time t+1 given action a. We estimated the transition matrix by counting the observed transitions in the development cohort and converting the counts to a stochastic matrix. To enhance safety, we limited the set of actions to frequently observed choices made by clinicians, excluding transitions with fewer than twenty occurrences. This approach ensures that the RL policy will learn from treatment options with high safety^23^.
- R(s’, s, a) is the immediate reward received for a transition. Transitions to desirable states yield positive rewards, while reaching undesirable states incurs penalties. In our model, if s’ is the final state and the patient experiences no CVD occurrence within one year, a high positive reward is given; conversely, a high negative reward is assigned if CVD occurs^23^. For patient actions involving LMD, a small penalty is applied to account for potential side effects^21^. The specific penalty values were determined through manual prioritization based on qualitative evaluation of the RL policy decision boundary during model development.
- γ is the discount factor, which accounts for the decreasing importance of future rewards compared to immediate rewards. The common practice of γ in healthcare applications typically ranges between 0.9 to 0.99^19,20,22,23^. We chose a γ value of 0.99, indicating that we assign nearly equal importance to late and early occurrences of rewards^19,23^.

After defining and calculating the components of MDP, we employed policy iteration^23,26^, an offline model-based dynamic programming algorithm in RL. This algorithm learns a state-action value function Q_π_, which quantifies the expected long-term reward of choosing an action in a given state, and a policy π that selects the action with the highest reward according to Q_π_^17^.

The policy iteration process began with a random policy and iteratively evaluated and improved it until convergence to an optimal solution^49^.

1. Policy evaluation on the expected reward of policy *V*^π^(*s*):

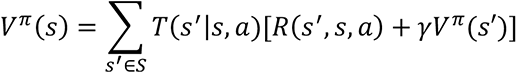

2. Policy improvement on the state-action value function *Q*^π^:

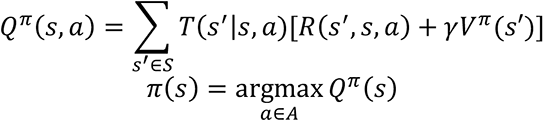

Until reaching the convergence of *π*(*s*).

### Model validation

We evaluated the policy value of the trained RL agent using a large independent validation time series dataset. To provide a comprehensive comparison, we introduced and evaluated two additional policies: the "no drug" policy, where the RL agent always chose not to prescribe any LMD, and the "random drug" policy, where the RL agent randomly selected an action from the available pool of actions. These policies served as baselines for comparison, allowing us to assess the performance of the RL agent against alternative decision-making strategies^23^.

**Calculation of policy value of clinicians’ policy.** We utilized our validation cohort C = [J_i_, i=1,2,…,n]. Each trajectory J_i_ = [(s_i_,t, a_i,t_, r_i_,t), t=1,2,…,τ_i_] represented a sequence of transitions (s_i,t_, a_i,t_, r_i_,t, s_i,t+1_) from step t to step t+1, where τ denotes the trajectory length. Within each trajectory, s_i,t_ represented the current state, a_i,t_ denoted the action taken, and r_i,t_ represented the immediate reward. The policy value of the clinicians’ policy is:

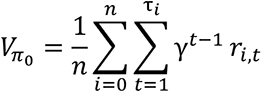

**Off-policy evaluation using importance sampling.** In order to ensure reliable estimates of the new policy’s performance before its deployment in real-world clinical settings, we engaged in off-policy evaluation (OPE)^17^. This process aimed to evaluate the RL policy’s performance using patient trajectories generated by the clinicians’ policy, as observed in the validation dataset.

Formally, within the context of OPE, we defined π0 as the behavior policy (the clinicians’ policy) and π1 as the RL policy. To account for the discrepancy between these two policies and estimate their policy value, we employed importance sampling^17,22^. Importance sampling is a widely recognized method in RL policy estimation, allowing us to correct for the differences between π0 and π1 and obtain accurate estimates of their respective policy values.

For trajectory i at time step t, the importance ratio is calculated as:

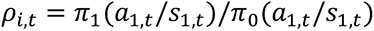

The weight of the trajectory is:

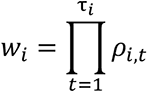

And the estimated value of the RL policy is:

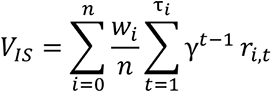

The same procedure was applied to the no drug policy and the random drug policy to estimate their policy value.

## Acknowledgements

Nil.

## Sources of funding

Nil.

## Disclosures

**E.Y.F.W.** has received research grants from the Health Bureau of the Government of the Hong Kong SAR, the Hong Kong Research Grants Council of the Government of the Hong Kong SAR, Narcotics Division, Security Bureau of the Government of the Hong Kong SAR, Social Welfare Department, Labour and Welfare Bureau of the Government of the Hong Kong SAR and National Natural Science Foundation of China, outside the submitted work. **F.T.T.L.** has been supported by the RGC Postdoctoral Fellowship under the Hong Kong Research Grants Council and has received research grants from the Health Bureau of the Government of the Hong Kong Special Administrative Region, outside the submitted work. **I.C.K.W.** reports grants from Amgen, Bristol-Myers Squibb, Pfizer, Janssen, Bayer, GSK and Novartis, the Hong Kong RGC, and the Hong Kong Health and Medical Research Fund in Hong Kong, National Institute for Health Research in England, European Commission, National Health and Medical Research Council in Australia, consulting fees from IQVIA and World Health Organization, payment for expert testimony for Appeal Court of Hong Kong and is a non-executive director of Jacobson Medical in Hong Kong and Therakind in England, outside of the submitted work. **C.L.C.** received research grants and the honorarium from Amgen, research grant support from HMRF, and the honorarium from Abbott. **C.S.L.C.** has received grants from the Food and Health Bureau of the Hong Kong Government, Hong Kong Research Grant Council, Hong Kong Innovation and Technology Commission, Pfizer, IQVIA, MSD, and Amgen; and personal fees from PrimeVigilance; outside the submitted work. **R.L.** has received grants from Hong Kong Research Grant Council, Hong Kong Innovation and Technology Commission; and research donations from Oxford Nanopore Technologies, outside the submitted work. All other authors declare no competing interests.

## Author contributions

Y.Z., R.L., and C.S.L.C. designed the research goals. Y.Z. and R.L. developed and evaluated the model. Y.Z., R.L., C.S.L.C., J.E.B., K.T., D.L., K.H.Y., F.T.T.L., E.Y.F.W., C.L.C., I.C.K.W. interpreted the results. Y.Z. drafted the manuscript. R.L. and C.S.L.C. supervised the project. All authors contributed to manuscript writing and revision.

## Ethics declarations

Ethical approval for this study was granted by the Institutional Review Board of The University of Hong Kong/HA Hong Kong West Cluster. Informed consent is waived by IRB in view of the retrospective nature of this study.

## Data availability

Sensitive patient data is not available. Restricted access for validation is available upon request. Please write to R.L. and C.S.L.C. for details.

---

**Extended Data Fig. 1.**
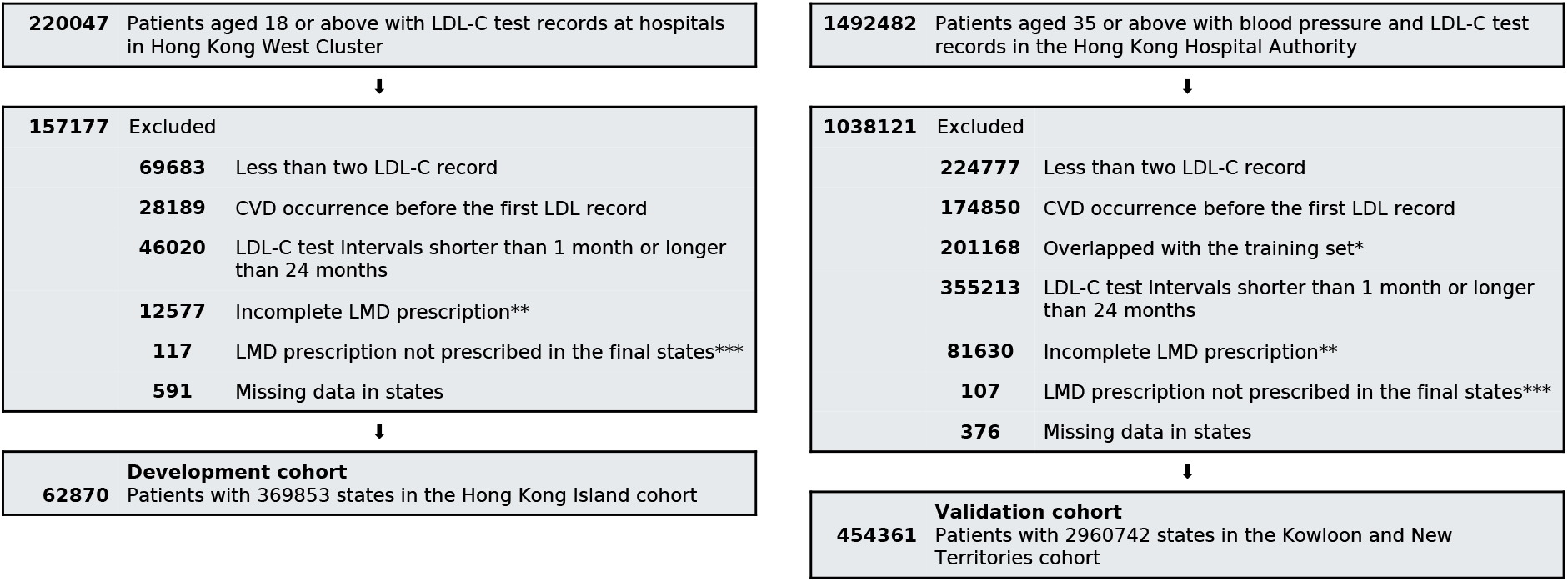
Selection of high quality and high confidence patient state-action trajectories. *With mostly frequently visited healthcare utilization at Hong Kong Island; Hong Kong West Cluster is a part of Hong Kong Island. **We consider a complete LMD treatment as the total duration of the prescription by one LMD to cover as least half of the interval of one LDL-C test. ***We consider final state prescription to be reliable representation of real clinician prescription. Prescription records in the middle may represent the change of treatment plans which are unstable and may represent false combinations of LMD, which should be restricted. CVD = cardiovascular disease. LDL-C = low-density lipoprotein cholesterol. LMD = lipid-modifying drug.

**Extended Data Fig. 2.**
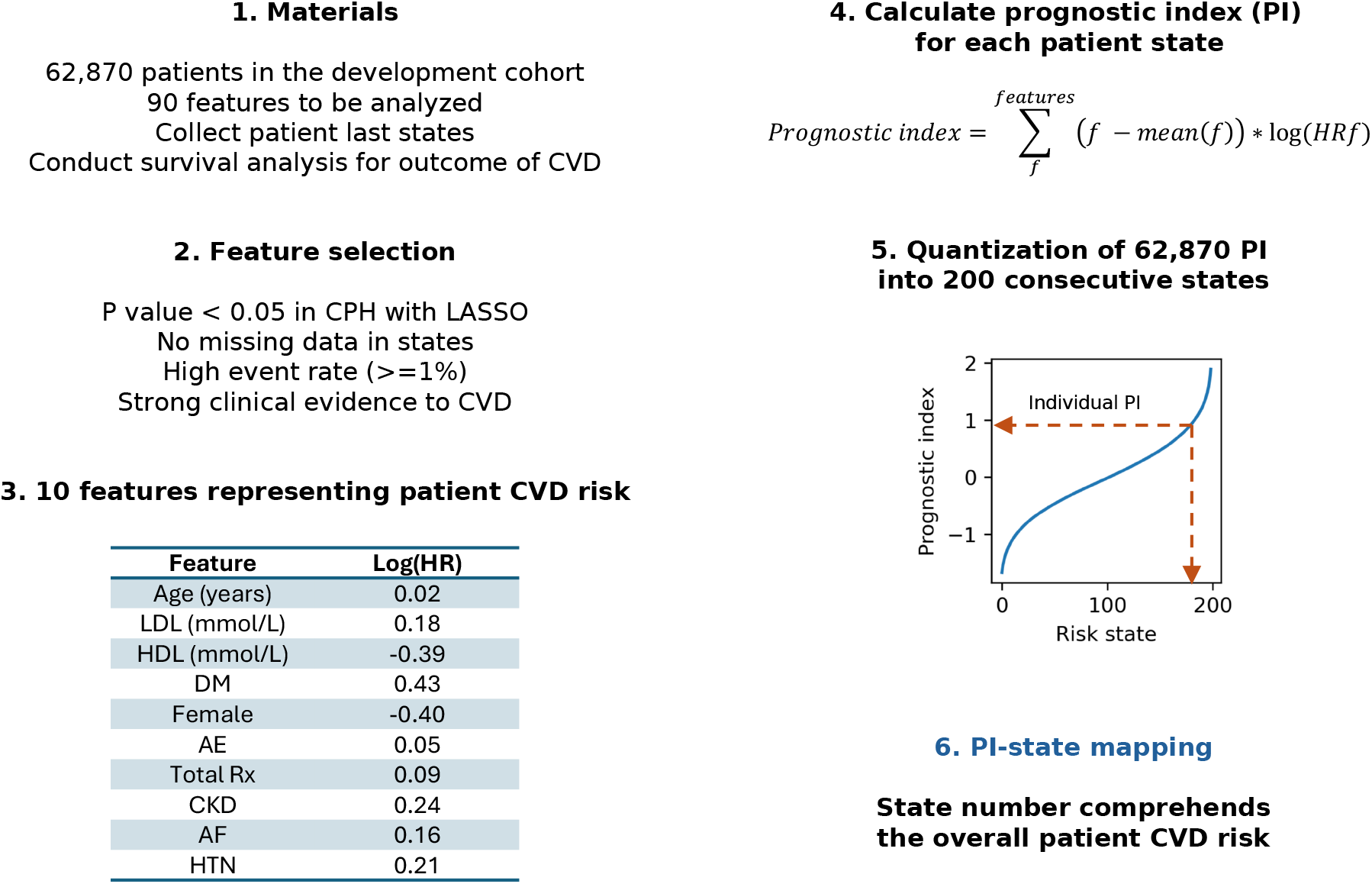
Risk-based state representation. CVD = cardiovascular disease. LASSO = least absolute shrinkage and selection operator. CPH = Cox proportional hazards model. LDL = low-density lipoprotein cholesterol. HDL = high-density lipoprotein cholesterol. DM = diabetes mellitus. AE = Accident and emergency visits within the last one year. Total Rx = concurrent medication records the number of drugs prescribed within one month. CKD = chronic kidney disease. AF = atrial fibrillation. HTN = hypertension.

**Extended Data Fig. 3.**
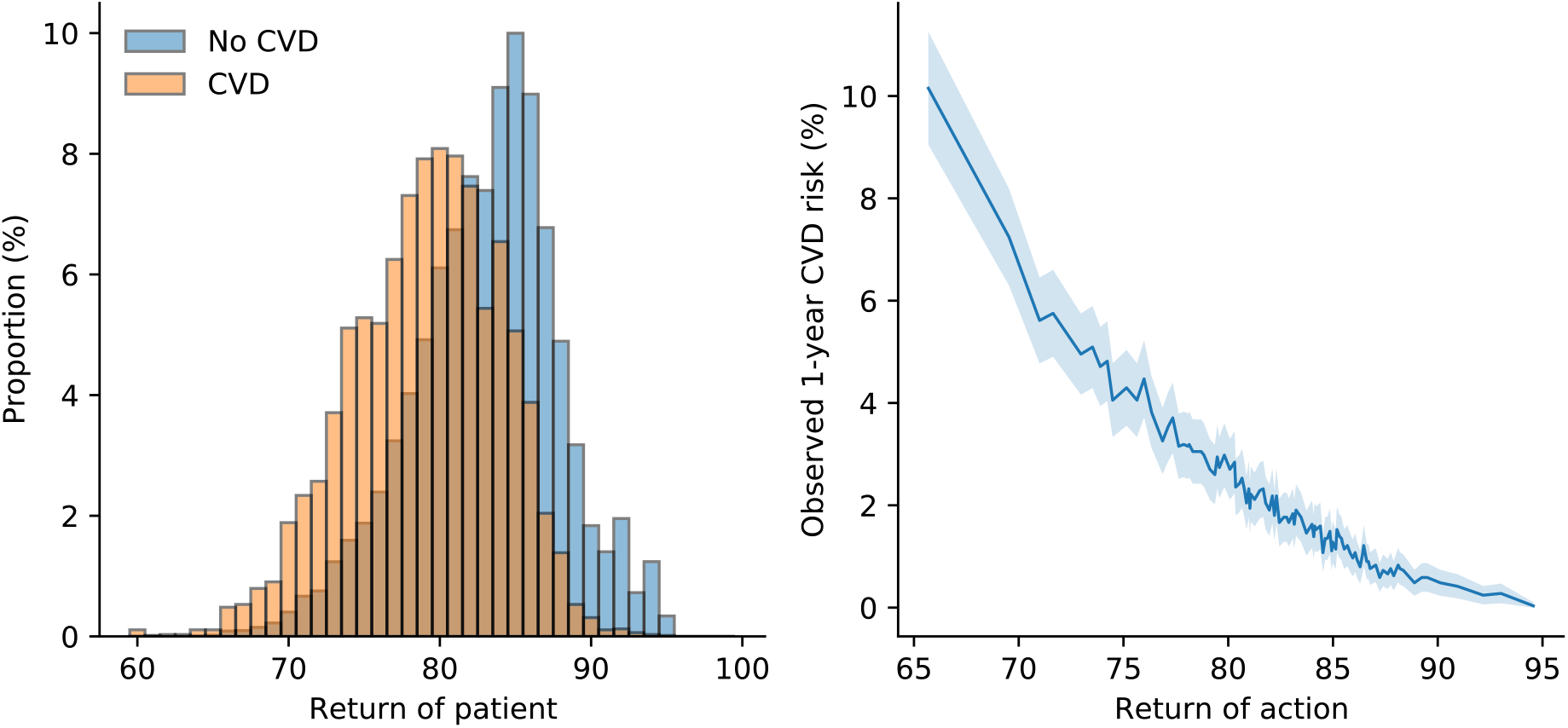
Model calibration check indicated the association between high-return actions and reduced CVD risk. (left) Bin plot showed the distribution of average return of patient actions within each trajectory comparing patients with observed 1-year CVD occurrence (tagged CVD) and those without (tagged No CVD) in the development cohort. (right) The relationship between the return of each action and patient 1-year CVD risk at the end of the trajectory in the development cohort. Return of actions were sorted into 100 bins, and the mean observed mortality was computed in each bin. The light color shaded blue area represents the standard error of the mean. Return of action was estimated using the fitted state action value function Q^π^, as illustrated in Methods.

**Extended Data Table 1.**
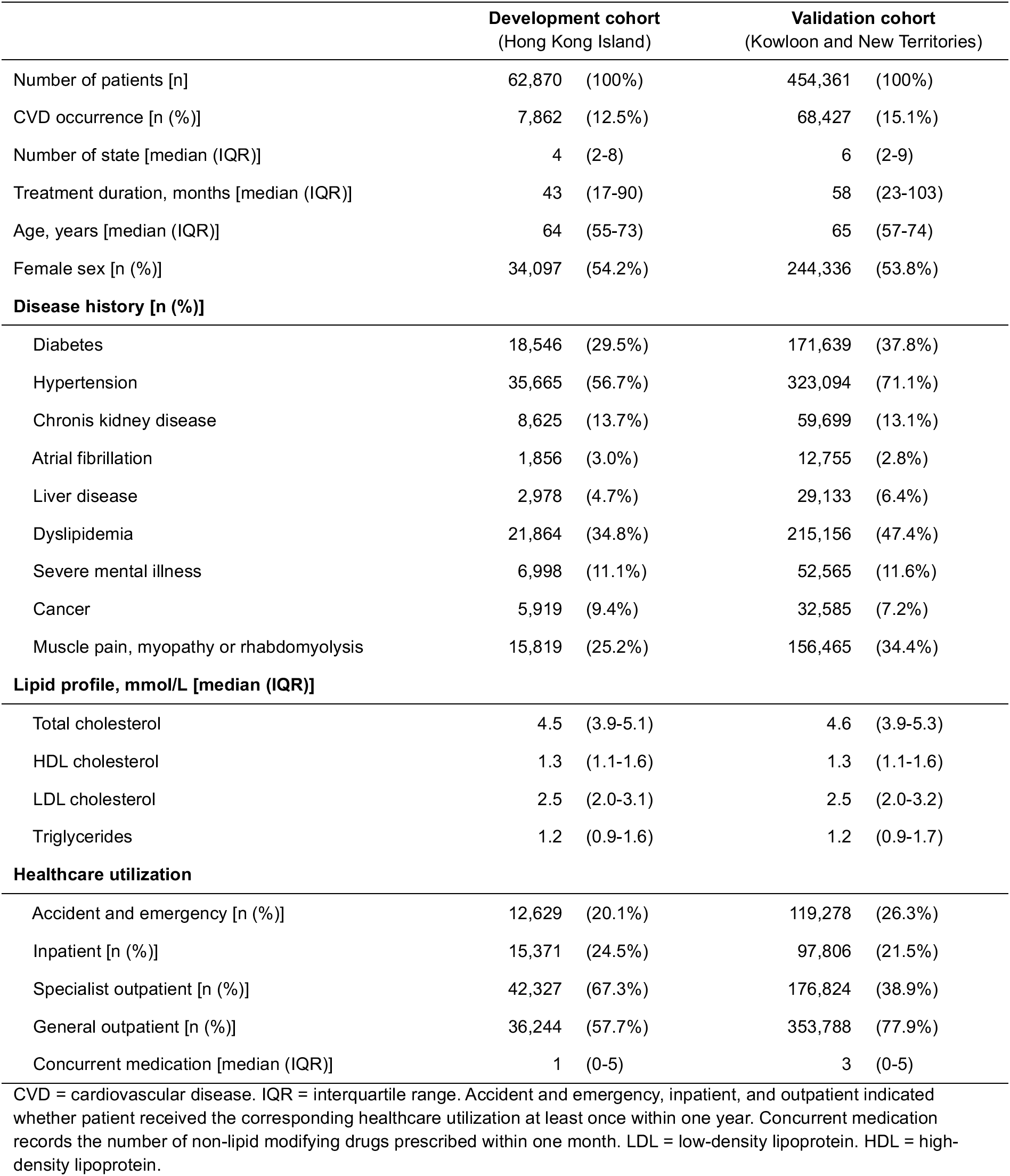
Description of datasets.

**Supplementary Table 1.** Summary of 90 variables in the development cohort Category (number) Covariates.

| Category (number) | Covariates |
| --- | --- |
| Disease history (41) | injury and poisoning, chronic obstructive pulmonary disease, arrhythmia and conduction disorders, Huntington's disease, Down's syndrome, cardioversion, liver disease, dialysis, hypothyroidism, history of dyslipidemia, renal disease, defibrillator insertion, heart transplantation, cardiomyopathy, rheumatoid arthritis, severe mental illness, diabetes, oxygen therapy/ventilator/intubation, systemic lupus erythematosus, erectile dysfunction, hypertension, cancer, Creutzfeldt-Jakob disease, revascularization, obesity, smoker, nephrotic syndrome, pacemaker implantation, dementia, major organ bleeding, muscle pain, myopathy or rhabdomyolysis, cardiac wall/valve/shunt replacement/repairment, memory loss, mild cognitive impairment, thyroid disease, alcohol user, chronic kidney disease, atrial fibrillation, Parkinson's disease, asthma, migraine |
| Demographics (2) | age, sex |
| Healthcare and medication count (5) | accident and emergency, inpatient, specialist outpatient, general outpatient, concurrent medication |
| Family disease history (2) | diabetes, cardiovascular disease |
| Laboratory tests (40) | albumin, alkaline phosphatase, alanine aminotransferase, aspartate aminotransferase, basophil, bicarbonate, bilirubin, calcium, creatine kinase, creatinine, c-reactive protein, diastolic blood pressure, estimated glomerular filtration rate, eosinophil, erythrocyte sedimentation rate, fasting glucose, hemoglobin, glycated hemoglobin, international normalized ratio, lymphocyte, monocyte, neutrophil, blood pH, platelet, partial pressure of oxygen, potassium, prothrombin time, red blood cell, systolic blood pressure, sodium, free thyroxine, troponin I, troponin T, thyroid stimulating hormone, white blood cell, total cholesterol, high-density lipoprotein cholesterol, low-density lipoprotein cholesterol, triglycerides, body mass index |

**Supplementary Table 2.** Results of feature selection using LASSO.

| Variables | P value | HR (95% CI) | Log(HR) (95% CI) |
| --- | --- | --- | --- |
| inpatient | 0.309 | 1.002 (0.998, 1.006) | 0.002 (-0.002, 0.006) |
| specialist outpatient | 0.000 | 1.022 (1.018, 1.026) | 0.022 (0.018, 0.026) |
| general outpatient | 0.995 | 1.000 (1.000, 1.000) | 0.000 (0.000, 0.000) |
| concurrent medication | 0.000 | 1.077 (1.071, 1.082) | 0.074 (0.069, 0.079) |
| injury and poisoning | 0.998 | 1.000 (1.000, 1.000) | 0.000 (0.000, 0.000) |
| chronic obstructive pulmonary disease | 0.999 | 1.000 (1.000, 1.000) | 0.000 (0.000, 0.000) |
| arrhythmia and conduction disorders | 0.996 | 1.000 (1.000, 1.000) | 0.000 (0.000, 0.000) |
| liver disease | 0.997 | 1.000 (1.000, 1.000) | 0.000 (0.000, 0.000) |
| dialysis | 0.000 | 1.642 (1.452, 1.857) | 0.496 (0.373, 0.619) |
| renal disease | 0.995 | 1.000 (1.000, 1.000) | 0.000 (0.000, 0.000) |
| rheumatoid arthritis | 0.995 | 1.000 (1.000, 1.000) | 0.000 (0.000, 0.000) |
| severe mental illness | 0.996 | 1.000 (1.000, 1.000) | 0.000 (0.000, 0.000) |
| oxygen therapy/ventilator/intubation | 1.000 | 1.000 (1.000, 1.000) | 0.000 (0.000, 0.000) |
| systemic lupus erythematosus | 0.998 | 1.000 (1.000, 1.000) | 0.000 (0.000, 0.000) |
| cancer | 0.995 | 1.000 (1.000, 1.000) | 0.000 (0.000, 0.000) |
| obesity | 0.997 | 1.000 (1.000, 1.000) | 0.000 (0.000, 0.000) |
| smoker | 0.999 | 1.000 (1.000, 1.000) | 0.000 (0.000, 0.000) |
| dementia | 0.995 | 1.000 (1.000, 1.000) | 0.000 (0.000, 0.000) |
| muscle pain, myopathy or rhabdomyolysis | 0.995 | 1.000 (1.000, 1.000) | 0.000 (0.000, 0.000) |
| thyroid disease | 0.998 | 1.000 (1.000, 1.000) | 0.000 (0.000, 0.000) |
| atrial fibrillation | 0.995 | 1.000 (1.000, 1.000) | 0.000 (0.000, 0.000) |
| asthma | 0.997 | 1.000 (1.000, 1.000) | 0.000 (0.000, 0.000) |
| low-density lipoprotein cholesterol | 0.000 | 1.057 (1.035, 1.079) | 0.055 (0.035, 0.076) |
| accident and emergency | 0.127 | 1.011 (0.997, 1.026) | 0.011 (-0.003, 0.026) |
| age | 0.000 | 1.023 (1.022, 1.024) | 0.023 (0.021, 0.024) |
| female | 0.000 | 0.768 (0.742, 0.794) | -0.264 (-0.299, -0.230) |
| chronic kidney disease | 0.000 | 1.105 (1.054, 1.158) | 0.100 (0.052, 0.147) |
| diabetes | 0.000 | 1.190 (1.137, 1.244) | 0.174 (0.129, 0.219) |
| hypertension | 0.000 | 1.096 (1.055, 1.138) | 0.091 (0.054, 0.129) |
| family history of diabetes | 0.000 | 1.506 (1.425, 1.592) | 0.410 (0.354, 0.465) |
| high-density lipoprotein cholesterol | 0.000 | 0.762 (0.730, 0.796) | -0.271 (-0.315, -0.228) |
| triglycerides | 0.995 | 1.000 (1.000, 1.000) | 0.000 (0.000, 0.000) |

Other 58 did not pass initial QC for LASSO because of either: 1) low event rate (<1%); 2) low data completeness. LASSO = least absolute shrinkage and selection operator.

**Supplementary Table 3.** Cox regression result on the 10 selected features.

| covariate | p value | HR (95% CI) | log(HR) (95% CI) |
| --- | --- | --- | --- |
| age | 0.000 | 1.023 (1.021, 1.024) | 0.022 (0.021, 0.024) |
| female | 0.000 | 0.671 (0.650, 0.694) | -0.399 (-0.432, -0.366) |
| high-density lipoprotein cholesterol | 0.000 | 0.677 (0.649, 0.707) | -0.390 (-0.433, -0.347) |
| low-density lipoprotein cholesterol | 0.000 | 1.191 (1.169, 1.214) | 0.175 (0.156, 0.194) |
| accident and emergency | 0.000 | 1.056 (1.046, 1.066) | 0.054 (0.045, 0.064) |
| concurrent medication | 0.000 | 1.094 (1.089, 1.098) | 0.089 (0.085, 0.094) |
| chronic kidney disease | 0.000 | 1.273 (1.221, 1.328) | 0.242 (0.200, 0.284) |
| atrial fibrillation | 0.000 | 1.169 (1.090, 1.253) | 0.156 (0.087, 0.226) |
| hypertension | 0.000 | 1.240 (1.197, 1.284) | 0.215 (0.179, 0.250) |
| diabetes | 0.000 | 1.537 (1.483, 1.593) | 0.430 (0.394, 0.466) |

**Supplementary Table 4.** Definition of cardiovascular disease.

| **Diagnosis** | **ICD-9-CM** |
| --- | --- |
| Peripheral artery disease | 440, 443.9 |
| Coronary heart disease | 410-414, 429.2, V45.81 |
| Stroke | 430-432, 433.01, 433.11, 433.21, 433.31, 433.81, 433.91, 434-436, 437.0, 437.1 |
| Congestive heart failure | 428 |

**Supplementary Table 5.** Definition of disease and symptoms.

| **Disease/Symptoms** | **ICD-9-CM** |
| --- | --- |
| Atrial fibrillation | 427.3 |
| Renal disease | 403.01, 403.11, 403.91, 404.02, 404.03, 404.12, 404.13, 404.92, 404.93, 580, 582, 583.0-583.7, 585-587, 588.0, 589, 590, 593.0-593.2, 593.6, 593.8, 593.9, 599.7, 753.0-753.4, 966.1, V42.0, V45.1, V56 |
| Chronic kidney disease | 585 |
| Dialysis | 585.9, V56.0, V56.8, 39.95 |
| Diabetes | 250 |
| Down's syndrome | 758.0 |
| Hypertension | 401-405 |
| Arrhythmia and conduction disorders | 426, 427 |
| Cardiomyopathy | 425 |
| Angina | 413 |
| Coronary artery bypass graft | 414.04, V45.81 |
| Myocardial infarction | 410 |
| Dyslipidemia | 272 |
| Thyroid disease | 240-244 |
| Liver disease | 570-573 |
| Migraine | 346 |
| Nephrotic syndrome | 581 |
| Rheumatoid arthritis | 446.5, 710.0-710.4, 714.0-714.3, 725 |
| Several mental illnesses | 290-319 |
| Systemic lupus erythematosus | 710.0 |
| Obesity | 278 |
| Dementia | 290, 291, 292.82, 294, 331 |
| Chronic obstructive pulmonary disease | 490-492, 494, 496 |
| Asthma | 493 |
| Alcohol use | 265.2, 291, 303, 305.0, 357.5, 425.5, 535.3, 571.0- 571.3, 980, V11.3 |
| Smoker | 305.1, V15.82, V15.83, 649.0 |
| Cancer | 140-209, 230-239 |
| Pacemaker implantation | 37.7, 37.8 |
| Defibrillator insertion | 37.94-37.98 |
| Cardioversion | 99.61 |
| Cardiac wall/valve/shunt replacement/repairment | 39.0-39.2 |
| Echocardiography | 37.28 |
| Heart transplantation | 37.51 |
| Oxygen therapy/ventilator/intubation | 00.49, 93.90, 96.01-96.05, 96.7 |
| Erectile dysfunction | 607.84 |
| Major organ bleeding | 578.0, 578.1 |
| Muscle pain, myopathy, or rhabdomyolysis | 728.8, 729.9, 791.3, 781.99 |
| Injury and poisoning | 800-989 |
| Parkinson's disease | 332 |
| Huntington's disease | 333.4 |
| Mild cognitive impairment | 331.83 |
| Memory loss | 780.93 |
| Creutzfeldt-Jakob disease | 046.1 |
| Hypothyroidism | 243-244 |

**Supplementary Table 6.** Definition of drugs.

| **Drug class** | **BNF chapter** |
| --- | --- |
| Corticosteroids | 1.5.2, 1.7.2, 3.2, 6.3, 8.2.2, 10.1.2, 11.4.1, 13.4 |
| H2-receptor antagonists | 1.3.1 |
| Proton-pump inhibitors | 1.3.5 |
| Anti-arrhythmic drugs | 2.3.2 |
| Psychotropic drugs | 4.1, 4.2, 4.3, 4.4 |
| Antihypertensive drugs | 2.2, 2.4, 2.5.1, 2.5.2, 2.5.4, 2.5.5, 2.6.2 |
| Anticoagulants | 2.8.1, 2.8.2 |
| Antiplatelet drugs | 2.9 |
| Antidiabetic drugs | 6.1.1.1, 6.1.1.2, 6.1.2.1, 6.1.2.2, 6.1.2.3 |
| Lipid-modifying drugs | 2.12 |
| Nicotine replacement therapy | 4.10.2 |
| Oestrogen | 6.4.1 |
| Testosterone | 6.4.2 |
| Non-steroidal anti-inflammatory drugs | 10.1.1 |
| Thyroid hormones | 6.2.1 |
| Antithyroid drugs | 6.2.2 |

## Notes

### Funding Statement

This study did not receive any funding.

### Author Declarations

Ethical approval for this study was granted by the Institutional Review Board of The University of Hong Kong/HA Hong Kong West Cluster

